# Evaluation of a process to implement advance care planning conversations in primary care: uptake and patient experience

**DOI:** 10.1101/2022.08.04.22278435

**Authors:** Neha Arora, Dale Guenter, Abe Hafid, Dawn Elston, Erin Gallagher, Samantha Winemaker, Nicolle Hansen, Heather Waters, Michelle Howard

## Abstract

**Objective:** Advance care planning (ACP) can support patients in achieving current and future medical care that aligns with their values and goals. In primary care, a lack of standardized processes hinders implementation of ACP conversations. This study reports a quality improvement process to identify and engage patients and clinicians in ACP.

**Methods:** Primary care clinicians received training in conversations based on the Serious Illness Conversation Guide and tools to support ACP. In December 2019, patients 65 years of age and older with chronic obstructive pulmonary disease were systematically identified, mailed ACP resources and telephoned by the clinic to invite them to an ACP appointment. We tracked the attendance of the patients and evaluated patient experience using a survey.

**Results:** Of the 91 patients telephoned, 50 were reached, and 27 attended the appointment. Further efforts were suspended in March 2020 due to the COVID-19 pandemic. Thirteen patients completed the survey. There were statistically significant increases in the patient’s perception of being heard and understood by their physician, feeling hopeful about quality of life and feeling peaceful.

**Conclusion:** This study provides evidence that with training, tools and processes, patients and primary care clinicians can be effectively engaged in ACP conversations.

**KEY MESSAGES:** *What was already known?:*

- Training resources exist for help clinicians enhance their advance care planning communication skills
- Besides a need for skills, other practical challenges exist in implementing advance care planning in family practice

*What are the new findings?:*

- Structured patient identification and preparation can facilitate advance care planning conversations in family practice
- Patients reported positive experiences of the conversations

*What is their significance?:*

- Clinical: It is important to move beyond clinician training alone to implement processes in family practice to trigger advance care planning conversations
- Research: Further research to identify effective scalable approaches to triggering and implementing advance care planning conversations in family practice would be beneficial

## BACKGROUND

Advance care planning (ACP) and communication about goals of care are important activities for people living with serious illness, to ensure current and future medical care aligns with patients’ values and goals.[1,2] Primary care clinicians tend to have long-standing relationships with patients, making this setting for the initiation of ACP. Despite the known benefits of ACP, [3] clinicians infrequently initiate conversations with patients about this topic.[4] Barriers commonly cited include their lack of specific skills, the absence of supporting tools and processes to integrate the activity into routine care and the perception that patients are unprepared for these conversations.[5]

The Serious Illness Conversation Guide (SICG) is a tool to improve clinicians’ skills for conversations about values and goals among patients with serious illness. Use of the guide has shown promising results in improving end-of-life outcomes in outpatient cancer care[3] and primary care settings.[6, 7] As part of an initiative to increase ACP in a large interprofessional teaching practice in Ontario, Canada, we previously provided SICP training to family physicians. After several months of regular follow-up, no ACP conversations were initiated. A questionnaire administered immediately after the training revealed that identifying and engaging patients were anticipated barriers.[8] This report presents the results of an evaluation of a quality improvement initiative to overcome the barriers, which involved identifying and preparing patients and family physicians for ACP conversations.

## METHODS

The quality improvement project to increase patient and physician engagement in ACP conversations, built on prior SICP training,[8] was undertaken from November 2019 to June 2020.

### Setting

The project took place at the McMaster Family Health Team (MFHT), comprised of approximately 40 family physicians and diverse allied health professionals at two family medicine clinics serving 40,000 patients in Hamilton, Ontario, Canada.

### Intervention

A detailed description of the intervention is provided in Supplementary File 1. Briefly, clinicians were trained in using the Serious Illness Conversation Guide as previously described.[8] They were given a “tip sheet” pocket card with guidance on facilitating GoC conversations based on key points from the training, including a suggested template for EMR documentation of the conversation. The initiative focused on patients with chronic obstructive pulmonary disease (COPD) as an exemplar population where it would be expected that conversations about care goals would occur at some point. A list of patients over 65 years with COPD was generated, and physicians identified patients who could benefit from the conversation. Clinic staff mailed information packages to patients. One to two weeks later, clinic staff telephoned patients to set up an appointment with the physician for those who agreed.

### Evaluation of Patient Experience

After the visit, physicians asked patients if they would complete an evaluation survey. If the patients consented to be contacted, a research assistant called them to administer a questionnaire by telephone. A retrospective pre-post (before and after the conversation) questionnaire was used, adapted from the SICP evaluation survey,[7] which explores the impact of the conversation (e.g., illness understanding, feeling of hopefulness, feeling heard and understood by the physician). We added questions on processes, such as how much the patient had thought about their values, wishes, and priorities and their confidence in talking to their substitute decision-maker and their physician(s) about their wishes. Responses were on 5-point Likert type scales (‘not at all’ to ‘a great deal’). Open-ended questions elicited additional perceived impacts of the conversation and suggestions to improve the process. Demographic characteristics of the participants (age, sex, education level, ethnicity, self-rated health and quality of life) were also collected.

### Analyses

Results were analyzed using descriptive statistics because the data were not normally distributed. We compared responses to questions as rated before and after the appointments using a Wilcoxon signed-rank test. Data was analyzed in IBM SPSS Statistics for Windows (Version 26.0. Armonk, NY: IBM Corp). The significance level was set at an alpha (two-sided) of less than 0.05. Patient comments were coded, and themes related to patients’ overall experience were extracted.

The project was approved by the Hamilton Integrated Research Ethics Board (HiREB #2018-4611).

## RESULTS

### Uptake of Goals of Care Conversations

In November 2019, 503 eligible patients were identified who were rostered with 37 physicians. Twenty-seven physicians actively participated in the process. From December 2019 to March 2020, 124 patients were mailed an invitation, and the clinic called 91 patients to set up an appointment (Figure 1). Clinic staff were able to speak with 50 patients, of whom 17 declined the appointment and 33 agreed to book an appointment. At this point, all research activity at the clinics was stopped due to COVID-19 pandemic restrictions. Ultimately, 27 patients attended the appointment before the survey was fully suspended due to ongoing COVID-19 restrictions.

### Patient Experience

Twenty-seven patients attended the appointment with their MRP. In some instances, a resident physician was also present. Thirteen patients (48%) completed the questionnaire, 6 patients (22%) declined, 3 patients (11%) withdrew, and 5 patients (19%) were not approached or were unreachable. Common reasons for patients declining the appointment included being busy, not interested, or having already planned for end of life. Some patients withdrew or cancelled their appointments due to the COVID 19 pandemic.

Patients had a mean age of 77 years. Of the 13 participants, 54 % were males, 84% had completed secondary education or higher, and 100% were white/European. Eighty-three percent of participants had never discussed this topic with their physicians before. All responses to the questions of how the conversation impacted patients were rated as higher following the GoC appointment. There were statistically significant differences in the patient’s perception of being heard and understood by their doctor (mean after 4.4, standard deviation [SD] 0.7 versus before 3.6 [SD 0.8]; p=.001), feeling hopeful about quality of life (mean after 4.1 [SD 0.9] versus before

[SD 1.0]; p=0.03), and feeling peaceful (mean after 4.2 [SD 0.8] versus before 3.6 [SD 0.7]; p=0.03). Patients found it worthwhile talking about these issues with their doctor (mean 4.5, SD 0.7).

Patients were also asked to provide feedback about the process and how this appointment made them feel. Major themes emerging from these answers indicated that patients appreciated the initiative and spent more time with their MRP. Patients also felt hopeful for the future and relieved because they were better informed. One patient noted: *“I do feel different to the point that I feel it is nice to have my values and preferences for the future on record. It’s nice to know to the family doctor also knows what I want*.*”*

## DISCUSSION

Our findings suggest that patient education and invitation to an ACP conversation by the family physician’s practice can activate some patients. Through proactive identification and outreach to patients, the clinics were able to engage patients with a serious illness to participate in conversations. Thirty-three out of 50 patients agreed to schedule an appointment, and most attended. Patient experience of the ACP conversation was generally positive.

Research has shown that patients wish for their healthcare provider to initiate ACP conversations;[9] however, we found that not all patients were interested at the time. Our participation rate was consistent with other studies inviting patients in primary care to engage in research on this topic.[10-12] Selecting patients who clinicians perceive as needing ACP and creating dedicated time can overcome barriers in clinical settings.

We used age, and the presence of one life-limiting condition as a simple criterion easily searched in the EMR to identify patients for ACP conversations. There is increasing interest in identifying patients who would benefit from an ACP conversation using automated mortality prediction algorithms. A review of rules that predict mortality between six months and five years among community-dwelling adults found ten algorithms that could be implemented in primary care, typically including age, sex, diagnoses and lifestyle risk factors.[13] Given that most patients with a chronic condition have a longitudinal relationship with their primary care provider, a combination of automated prompts and patient-centred approaches may be ideal.

The processes undertaken to engage physicians and patients may seem resource-intensive for clinics to undertake on an ongoing basis. However, these efforts may result in greater uptake over time if patients who initially declined appointment invitations raise the issue spontaneously at a future visit. Patients who accepted the appointment provided positive feedback and may have been already prepared and interested. Unfortunately, further follow-up contacts and appointments were not possible starting March 2020 due to the onset of the global COVID-19 pandemic, which resulted in the cessation of much research and re-prioritizing of clinic activities. Future studies should assess improvement initiatives over a longer period to determine which are successful for primary care clinics to incorporate ACP conversations as part of routine care.

## CONCLUSION

Patient-related barriers to ACP conversations have long been discussed in the literature. Still, this study provides evidence that patients can be engaged in ACP conversations in primary care with the right tools and processes. A process to implement ACP conversations in primary care using structured tools and approaches appeared to be successful, and patients reported positive experiences.

**Table 1:**
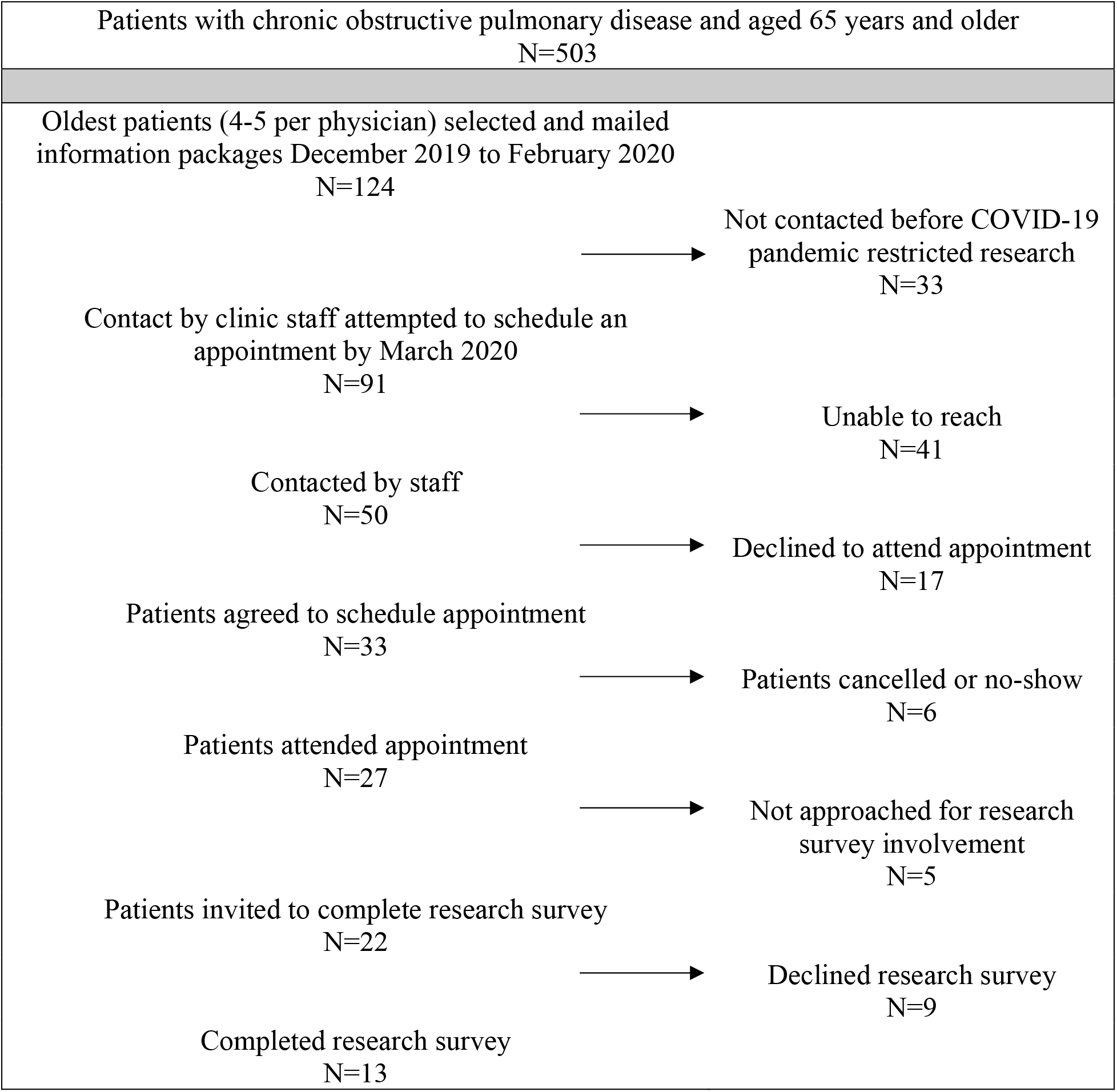
Flow of patients through the study

| Patients with chronic obstructive pulmonary disease and aged 65 years and older<br>N=503 |  |
| --- | --- |
| Oldest patients (4-5 per physician) selected and mailed information packages December 2019 to February 2020<br>N=124 |  |
| → | Not contacted before COVID-19 pandemic restricted research<br>N=33 |
| Contact by clinic staff attempted to schedule an appointment by March 2020<br>N=91 |  |
| → | Unable to reach<br>N=41 |
| Contacted by staff<br>N=50 |  |
| → | Declined to attend appointment<br>N=17 |
| Patients agreed to schedule appointment<br>N=33 |  |
| → | Patients cancelled or no-show<br>N=6 |
| Patients attended appointment<br>N=27 |  |
| → | Not approached for research survey involvement<br>N=5 |
| Patients invited to complete research survey<br>N=22 |  |
| → | Declined research survey<br>N=9 |
| Completed research survey<br>N=13 |  |

## Data Availability

All data produced in the present study are available upon reasonable request to the authors

## Contributorship Statement

DG and MH designed the study, NA, AH, DE contributed to implementing the study and acquisition of the data, NA analyzed the data, NA, DG, AH, DE, EG, SW, NH, HW and MH contributed to the interpretation of the data, NA wrote the manuscript, DG, AH, DE, EG, SW, NH, HW and MH critically revised the manuscript and NA, DG, AH, DE, EG, SW, NH, HW and MH gave final approval of the version to be published.

## REFERENCES

1. Canadian Hospice Palliative Care Association. Quality End-of-Life Care Coalition of Canada Progress Report: Blueprint for Action 2020-2025. Ottawa, Ontario: 2020. Retrieved from https://www.chpca.ca/news/the-quality-end-of-life-care-coalition-of-canada-qelccc-releases-the-blueprint-for-action-2020-2025/. Accessed June 24, 2022.

2. You JJ, Dodek P, Lamontagne F, Downar J, et al. What really matters in end-of-life discussions? Perspectives of patients in hospital with serious illness and their families. CMAJ 2014;186:E679–E87.

3. Bernacki R, Paladino J, Neville BA, et al. Effect of the serious illness care program in outpatient oncology: a cluster randomized clinical trial. JAMA Intern Med 2019;179:751–9.

4. Howard M, Langevin J, Bernard C, et al. Primary care clinicians’ confidence, willingness participation and perceptions of roles in advance care planning discussions with patients: a multi-site survey. Fam Pract 2020;37:219–26.

5. Howard M, Bernard C, Klein D, et al. Barriers to and enablers of advance care planning with patients in primary care: survey of health care providers. Can Fam Physician 2018;64:e190–8.

6. Lakin JR, Koritsanszky LA, Cunningham R, et al. A systematic intervention to improve serious illness communication in primary care. Health Aff 2017;36:1258–64.

7. Lakin JR, Neal BJ, Maloney FL, et al. A systematic intervention to improve serious illness communication in primary care: Effect on expenses at the end of life. Healthc 2020;8:100431. doi: 10.1016/j.hjdsi.2020.100431 [published Online First: 14 May 2020]

8. Hafid A, Howard M, Guenter D, et al. Advance care planning conversations in primary care: a quality improvement project using the Serious Illness Care Program. BMC Palliat Care 2021;20:122. doi:10.1186/s12904-021-00817-z

9. Hall A, Rowland C, Grande G. How should end-of-life advance care planning discussions be implemented according to patients and informal carers? A qualitative review of reviews. J Pain Symptom Manage 2019;58:311–35.

10. Sudore RL, Schillinger D, Katen MT et al. Engaging diverse English-and Spanish-speaking older adults in advance care planning: the PREPARE randomized clinical trial. JAMA Intern Med 2018;178:1616–25.

11. Heyland DK, Heyland R, Bailey A, Howard M. A novel decision aid to help plan for serious illness: a multisite randomized trial. CMAJ Open. 2020 Apr 1;8(2):E289–96.

12. Howard M, Elston D, Borhan S, et al. Randomised trial of a serious illness decision aid (Plan Well Guide) for patients and their substitute decision-makers to improve engagement in advance care planning. BMJ Support Palliat Care 2022;12:99–106.

13. Kim P, Daly JM, Berry-Stoelzle MA, et al. Prognostic indices for advance care planning in primary care: a scoping review. J Am Board Fam Med 2020;33:322–38.

