## Supplemental Table 1 for "Evaluation of a process to implement advance care planning conversations in primary care: uptake and patient experience"

**Supplementary File 1: Description of the Advance Care Planning (ACP) initiative in family practice**

| **Steps** | **Description** |
|---|---|
| **1. Clinician training** | Physicians participated in training on the Serious Illness Conversation Guide (SICG), which included a half-day workshop comprised of didactic teaching and role-playing using the conversation Serious Illness Conversation Guide with trained standardized patients. An additional six physicians attended a later one-hour didactic in-service on person-centred decision making, which contained concepts overlapping with the SICG training while also including training on the legal framework in the province (https://www.speakupontario.ca/e-learning-module/). |
| **2. Patient identification** | Without a systematic method to identify patients at the highest risk for health decline and death in primary care, we focused this initiative on patients with chronic obstructive pulmonary disease (COPD). This population with a progressive life-limiting illness is relatively common in primary care and exemplifies a starting place for routinizing ACP conversations in primary care.<br><br>Using the disease registry created from the electronic medical record, a list of patients over 65 years of age with a diagnosis of COPD was generated for each participating physician. Participating physicians were presented with their respective patient lists and asked to identify patients they believed would benefit most from an ACP conversation. |
| **3. Patient communication** | Administrative staff at clinics mailed information packages to identified patients between December 2019 and January 2020. The packages included an explanation letter from their physician and a Speak Up! Ontario Advance Care Planning workbook (https://www.speakupontario.ca/resource/acp-workbook-en/). This ensured that all patients received background information on the topic, irrespective of participation. |
| **4. Physician guides** | To assist with communication of the project to providers, we created a brochure describing: 1) the initiative; 2) which patients would be invited; and 3) how the patient would be invited, including a copy of the letter to patients. This was provided to all physicians 2-4 weeks before patients were contacted and invited to appointments. The brochure also included a link to the online modules for learning about goals of care conversations.<br><br>We also created a "tip sheet" pocket card with guidance on facilitating ACP conversations based on key points from both training resources. The tip sheet included a suggested template for documentation and a fictitious example of a documented conversation, but no permanent changes were made to the EMR. |
| **5. Advance care planning appointment** | After approximately 7-14 days, clinic staff called each patient to book an ACP appointment with their respective most responsible physician. |

| | |
|---|---|
| | Each physician could choose to have a mentor attend the appointment if preferred. Mentors were clinicians (ten physicians and one nurse practitioner) who attended SICG training previously and had specific training and expertise in palliative care. Residents were also encouraged to attend the appointment. |
